# Molecular epidemiology of AY.28 and AY.104 delta sub-lineages in Sri Lanka

**DOI:** 10.1101/2022.02.05.22270436

**Authors:** Diyanath Ranasinghe, Deshni Jayathilaka, Chandima Jeewandara, Dumni Gunasinghe, Dinuka Ariyaratne, Tibutius Thanesh Pramanayagam Jayadas, Heshan Kuruppu, Ayesha Wijesinghe, Farha Bary, Deshan Madusanka, Pradeep Darshana, Dinuka Guruge, Ruwan Wijayamuni, Graham S. Ogg, Gathsaurie Neelika Malavige

## Abstract

**Background:** The worst SARS-CoV-2 outbreak in Sri Lanka was due to the two Sri Lankan delta sub-lineages AY.28 and AY.104. We proceeded to further characterize the mutations and clinical disease severity of these two sub-lineages.

**Methods:** 705 delta SARS-CoV-2 genomes sequenced by our laboratory from mid-May to November 2021 using Illumina and Oxford Nanopore were included in the analysis. The clinical disease severity of 440/705 individuals were further analyzed to determine if infection with either AY.28 or AY.104 was associated with more severe disease. Sub-genomic RNA (sg-RNA) expression was analyzed using periscope.

**Results:** AY.28 was the dominant variant throughout the outbreak, accounting for 67.7% of infections during the peak of the outbreak. AY.28 had three lineage defining mutations in the spike protein: A222V (92.80%), A701S (88.06%), and A1078S (92.04%) and seven in the ORF1a: R24C, K634N, P1640L, A2994V, A3209V, V3718A, and T3750I. AY.104 was characterized by the high prevalence of T95I (90.81%) and T572L (65.01%) mutations in the spike protein and by the absence of P1640L (94.28%) in ORF1a with the presence of A1918V (98.58%) mutation. The mean sgRNA expression levels of ORF6 in AY.28 were significantly higher compared to AY.104 (p < 0.0001) and B.1.617.2 (p < 0.01). Also, ORF3a showed significantly higher sgRNA expression in AY.28 compared to AY.104 (p < 0.0001). There was no difference in the clinical disease severity or duration of hospitalization in individuals infected with these sub lineages.

**Conclusions:** Therefore, AY.28 appears to have a fitness advantage over the parental delta variant (B.1.617.2) and AY.104 possibly due to the A222V mutation. AY.28 also had a higher expression of sg-RNA compared to other sub-lineages. The clinical implications of these should be further investigated.

## Introduction

The SARS-CoV-2 virus continues to result in outbreaks in many geographical regions, with the number of cases exponentially increasing in many countries due to the rapid transmission of Omicron ^1^. Of the five variants of concern (VOCs) that have been identified so far, the delta variant is associated with more severe disease compared to other variants^2,3^. Until Omicron emerged, the delta variant was the most transmissible variant and rapidly displaced all other VOCs and variants of interest^4^. Due to the higher transmissibility and increased virulence of the delta variant, outbreaks due to the delta wave were associated with the highest mortality, intensive care admissions and hospitalizations so far in all countries^1^.

As the delta variant was the dominant variant globally before emergence of Omicron for the longest time period during the COVID-19 pandemic, it gave rise to over 100 sub lineages. Phylogenetic Assignment of Named Global Outbreak (PANGO) nomenclature has currently assigned 122 sub lineages of delta (AY.1 – AY.122) that are distributed in different geographical regions^4^. The sub-lineages of the delta variants have not shown to be functionally different to the parental delta variant (B.1.617.2) and have shown to have a similar susceptibility to neutralizing antibodies^5^. However, some of these sub lineages such AY.4.2 have been assigned as a variant of interest due to its possible higher transmissibility compared to B.1.617.2 the parental delta variant^6^. While certain mutations such as the presence of A222V has shown to cause slightly higher viral titres, which is thought to result in a higher transmissibility^7^, mutations such as the E484K have a possibility of enhanced immune evasion^8^. Therefore, it is important to study the evolution and spread of different delta sub lineages to understand their transmission and to detect possible changes associated with virulence and immune evasion.

The largest Sri Lankan SARS-CoV-2 outbreak was due to the delta variant and its sub lineages from July to end of October 2021^9^. During the peak of this ‘delta wave’ the PCR positivity rates rose above 30% with case fatality rates reaching 6.35%^9^. Apart from the delta parental lineage B.1.617.2, two other delta sub lineages AY.28 and AY.104 were the predominant variants observed during this time period^4^. The AY.28 and AY.104 were assigned as Sri Lanka delta sub-lineages as they were found to originate in Sri Lanka and to be transmitted to all continents in the world^10,11^. In this study, we discuss lineage defining mutations, sub genomic RNA expression, relative frequency over time and clinical disease severity of individuals infected with either AY.28, AY.104 or B.1.617.2 in Sri Lanka.

## Methods

### Identification of AY.28 and AY.104 lineages in Sri Lanka

705 delta SARS-CoV-2 genomes sequenced by our laboratory from mid-May to November 2021 using Illumina and Oxford Nanopore were included in the analysis. Details of RNA extraction, library preparation, and analysis are given in supplementary methods. Of these 335/705 (47.5%) were assigned to AY.28 and 217/705 (28.5%) were assigned to AY.104, while 68/705 (9.6%) were assigned to B.1.617.2. The rest of the sequences were assigned to various AY sub-lineages of Delta (<24) by pangolin v3.1.16 (https://github.com/cov-lineages/pangolin) with the pangoLearn model released on 2021-11-25. In order to analyze the frequency of infections caused by these sub-lineages over time, we analyzed the relative change in the frequencies of AY.28 and AY.104 in the Colombo district from 15^th^ July to 30^th^ November 2021. As the frequency of delta was <50% of the SARS-CoV-2 viruses that were sequenced before 15^th^ July 2021, they were not included in the analysis. Metadata and statistical analysis of all the 705 samples are included in the supplementary table 1.

The clinical disease severity and vaccination status of 440/705 individuals who were found to be infected with the delta variant during this time period were further analyzed in order to determine if infection with either AY.28 or AY.104 was associated with more severe disease or with the type of vaccination. Those who were not hospitalized or who were hospitalized and were not given oxygen were considered as having mild illness, whereas those who were given oxygen or required intensive care admission were classified as moderate/severe based on the WHO guidelines in COVID-19 clinical disease classification ^12^. Pearson’s Chi-squared test was used to determine the associations between categorical variables (gender, vaccination, vaccine dose, and disease severity) and sub-lineages while pairwise T-tests with Bonferroni correction to compare means of age and hospitalization period. All statistical tests were done using R version 4.1.2.

### Mutational and phylogenetic analysis of AY.28 and AY.104

In addition to the delta variants sequenced by us, all the AY.28 (n=519) and AY.104 (n=493) sequences available at GISAID were downloaded from the GISAID database and aligned to the reference sequence using Nextalign (https://github.com/neherlab/nextalign). Amino acid mutations and their frequencies for Spike, ORF1a, ORF1b, and N proteins were calculated, and frequencies of ambiguous amino acids derived from ambiguous nucleotides were removed using in-house python scripts. The phylogenetic tree was inferred by Maximum Likelihood in IQ-Tree (version 1.6.12) using the GTR+G model of nucleotide substitution and 1000 replicates of ultrafast bootstrapping (-B 1000) and SH-aLRT branch test (-alrt 1000). The ML tree was then time stamped with TreeTime ^13^ (version 0.7.5) using least-squares criteria and the evolutionary rate of 1.1*10^−3^ subs/site/year as described by Duchene et a^14^. Six sequenced with inconsistent temporal signal were removed from the analysis. The tree was rendered using ggtree in R version 4.1.2.

### Sub-genomic RNA expression of AY.28 and AY.104

Sub-genomic RNA expression of 705 delta genomes (335 AY.28, 217 AY.104 and 68 B.1.617.2) were analyzed using periscope (https://github.com/sheffield-bioinformatics-core/periscope). This algorithm aligns raw reads against the SARS-CoV-2 reference genome (MN908947.3) and identifies reads that contain the leader sequence at their start position. Depending on the amplicon position, the sgRNA detected reads were counted and classified into each ORF of the virus. Means of sgRNA counts normalized to per 1000 genomic RNA reads, were then compared individually for each of the ORFs using the unpaired Wilcoxon test by adjusting p-values with the Holm method. Statistical analysis results are presented in supplementary table 2.

### Structural analysis of AY.28 and AY.104

The PyMOL Molecular Graphics System v.2.4.0 (https://github.com/schrodinger/pymol-open-source/releases) was used to map the location of the mutations defining the AY.28 and AY.104 onto the closed-conformation spike protein (PDB: 6ZGE).

## Results

We have previously described the changes in the SARS-CoV-2 variants from the onset of the pandemic to May 2021 in Sri Lanka ^15^. The first delta variant was identified in Sri Lanka on 22^nd^ May (AY.28), and the relative frequency of delta and sub lineages in Sri Lanka is shown in Figure 1A. By the first week of December 2021, out of all the delta variants sequenced from Sri Lanka, 481/974 (49.4%) belonged to AY.28 lineage, while only 320/974 (32.9%) were AY.104. Only 140/974 (14.4%) belonged to the parental delta variant, B.1.617.2, while 33/974 (3.39%) belonged to the AY.95 delta sub lineage that is thought to have originated in Maldives.

**Figure 1:**
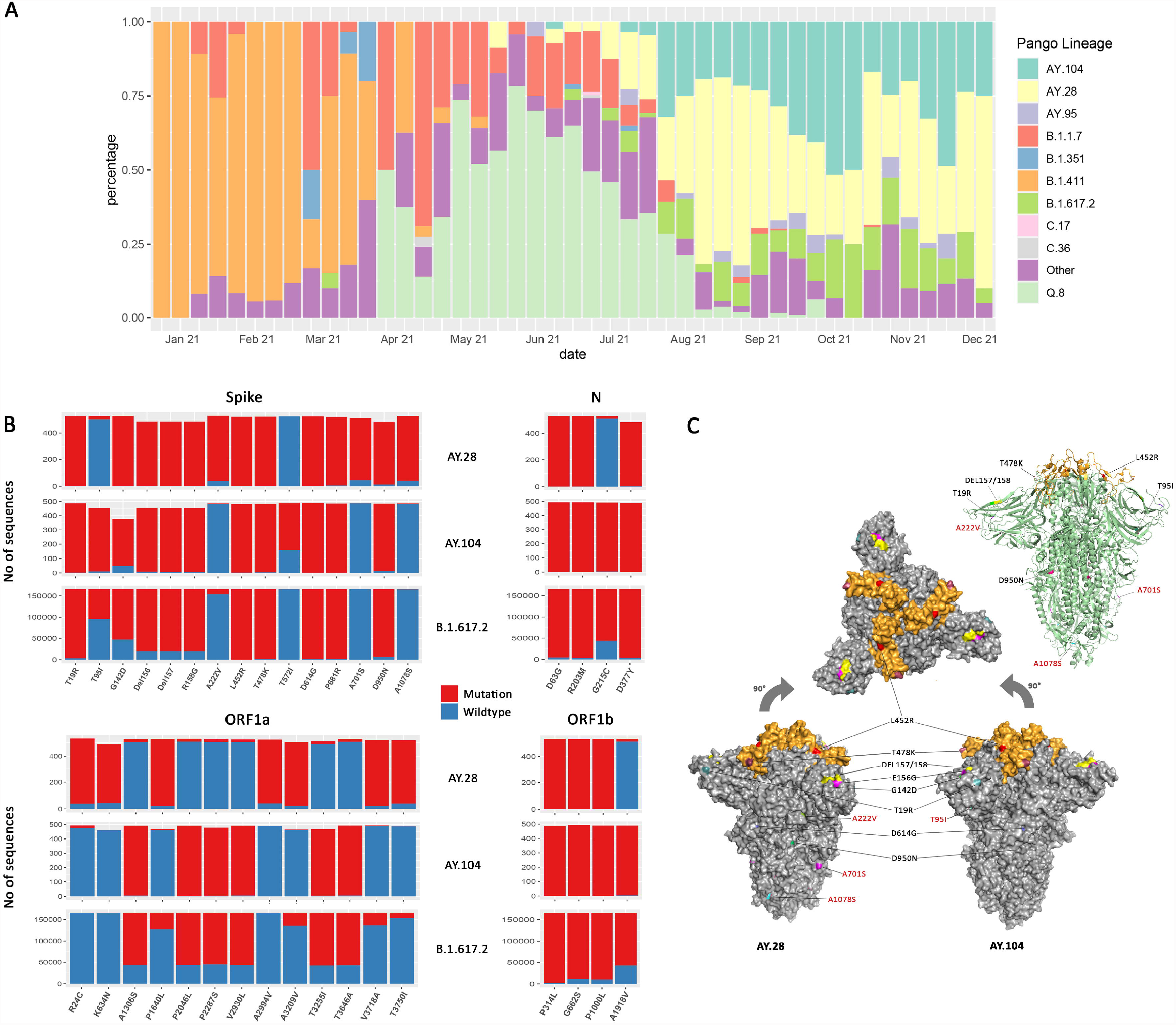
The mutation and structural changes of AY.28 and AY.104 sequences in comparison with B.1.617.2 (Delta) lineage. Weekly lineage frequencies of the sequenced cases in Sri Lanka since January 2021 were retrieved from the GISAID EpiCoV database (A). The frequency of mutations in the spike, ORF1a, ORF1b, and N proteins of AY.28 and AY.104 compared to those of the parental B.1.617.2 (Delta) lineage. The bar height represents total unambiguous amino acids at each position, mutated (red) and wildtype (blue) based on the analysis of the global sequences deposited in the EpiCoV database (B). The structural modelling of the pre-fusion surface representation of the SARS-COV-2 B.1.671.2 spike trimer (PDB: 6ZGE) and the mutations in the structure in AY.28 and AY.104 are shown. The receptor-binding domain (RBD) is colored in orange while the N-terminal and S2 subunit are shown in grey. Amino acid substitutions and deletions are colored on the surface and labels of the mutations specific for AY.28 and AY.104 are colored in red. On the right, a ribbon diagram is shown of the same.

### Mutational and structural analysis of AY.28 and AY.104

AY.28 had three lineage defining mutations in the spike protein: A222V (92.80%), A701S (88.06%), and A1078S (92.04%). ORF1a of AY.28 consists of seven lineage defining mutations: R24C, K634N, P1640L, A2994V, A3209V, V3718A, and T3750I with prevalence between 97.83% to 87.96%, and A1918 (96.59%) in ORF1b, and G215 (96.21%) in the N protein, which is present in the SARS-CoV-2 wild-type virus (Figure 1B). AY.104 was characterized by the high prevalence of T95I (90.81%) mutation and T572L (65.01%) mutation in the spike protein, A1918V (98.58%) in ORF1a, G215C mutation (98.98%) in N protein. In addition, it is characterized by the absence of P1640L (94.28%) in ORF1a as seen in the wild-type virus (Figure 1B).

The pre-fusion (closed-conformation) surface representation of the SARS-COV-2 B.1.671.2 spike trimer (PDB: 6ZGE) mapped by PyMOL v.2.4.0 is shown in figure 1C. The receptor-binding domain (RBD) is colored in orange while the N-terminal and S2 subunit are shown in grey. Amino acid substitutions and deletions that correspond to each delta sub-lineage are shown in two separate diagrams. Ribbon diagram view of the spike trimer common to both AY.28 and AY.104 of the same is shown on the top right (Figure 1C).

### Changes in the relative frequency of B.1.617.2 and the two delta sub lineages over time

In order to understand the possible transmissibility of each of the delta sub lineages in comparison to each other, we assessed the relative frequency of B.1.617.2 and the two sub-lineages of delta (AY.28 and AY.104), in the Colombo district. We could not extend this to an island wide analysis, as samples were sequenced infrequently from other districts. Of the 705 delta variants sequenced by us from 15^th^ July to 18^th^ October 440 were from the Colombo district. From July 2021 to October 2021, AY.28 accounted for 251/440 (57%) of the delta genomes, while AY.104 accounted for 109/440 (24.8%) and B.1.617.2 accounted for 39/440 (8%) of the genomes sequenced from the Colombo district. The changes in the frequency of the B.1617.2, AY.28 and AY.104 sequenced from the Colombo district are shown in figure 2.

**Figure 2:**
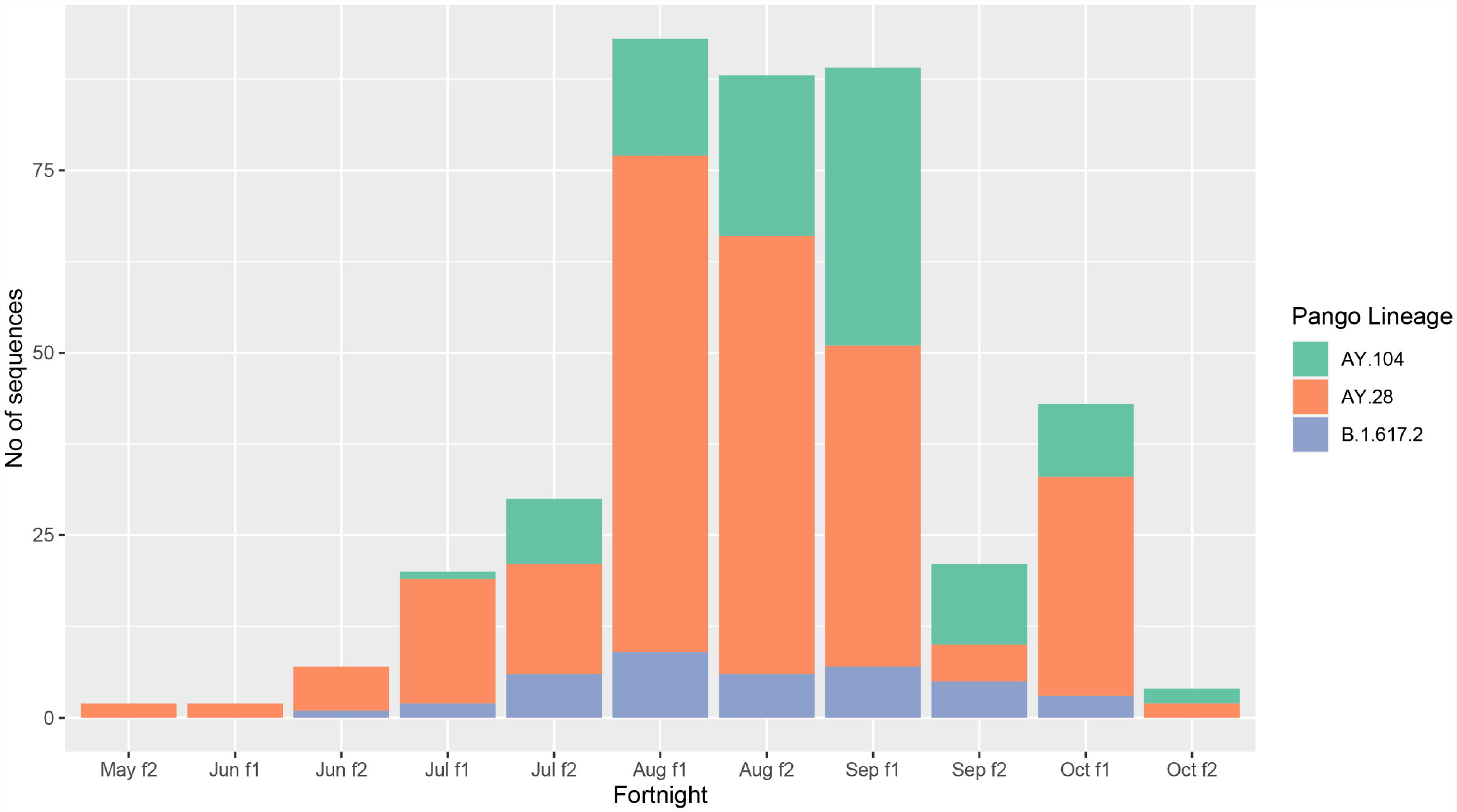
Prevalence of AY.28 and AY.104 against the original Delta (B.1.617.2) in Colombo district. Prevalence of main delta sub-lineages (AY.28, AY.104 and B.1.617.2) sequenced in Colombo district (N=440) since mid-May to November 2021 is shown for each fortnight, month and week.

The time resolved maximum likelihood tree of global AY.28 and AY.104 sequences compared to the parent B.1.617.2 sequences reported in Sri Lanka suggests the time of the most recent common ancestor (tMRCA) of AY.28 emerged around 20^th^ of January 2021. According to the analysis of tMRCA, AY.104 appears to have originated around 20^th^ April 2021. Interestingly, a separate node of AY.28 predominantly sampled in USA appears to have originated in mid-January 2021. The tips are color annotated according to the country where the sequences were identified, which shows Sri Lanka as the likely country of origin (Figure 3). Majority of the AY.28 and AY.104 sequences detected outside of Sri Lanka were reported from the United Kingdom, India, Japan, Canada and Australia (Figure 3).

**Figure 3:**
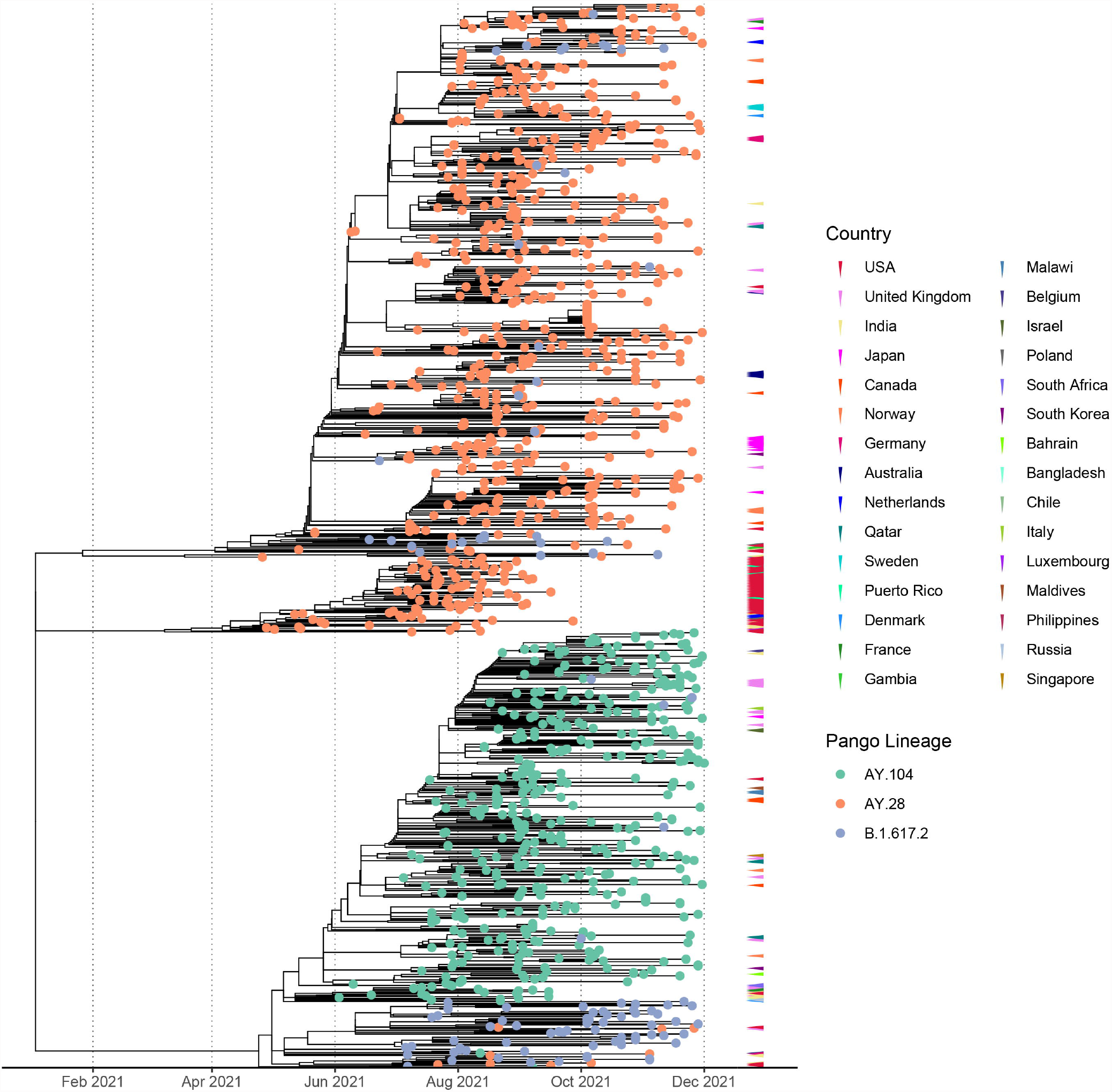
Divergence times of the phylogenetic trees of the global AY.28 and AY.108 viruses using molecular clock. Time-resolved maximum-likelihood phylogenetic tree of AY.28 and AY.104 cases sequenced globally and B.1.617.2 sequences only originating from Sri Lanka. The molecular clock was inferred using TreeTime with an evolutionary rate of 1.1 × 10-3 substitutions/site/year with a standard deviation of 0.00004. Non-local cases of AY.28 and AY.104 are color-coded according to the originating country on the outer layer.

### Sub-genomic RNA expression of Delta sub-lineages

We analysed 335 AY.28 genomes and 217 AY.104 and 68 B.1.617.2 for expression of sgRNA. The highest sgRNA expression was observed in the spike protein of B.1.617.2 and the sub-lineages followed by ORF7a (Figure 4). The mean sgRNA expression levels of ORF6 in AY.28 was significantly higher compared to AY.104 (p < 0.0001) and B.1.617.2 (p < 0.01). Also, ORF3a showed significantly higher sgRNA expression in AY.28 compared to AY.104 (p < 0.0001). However, AY.104 had significantly higher sgRNA expression in ORF8 compared to AY.28 (p < 0.0001), and B.1.617.2 (p < 0.0001). Also, the mean sgRNA expression of the nucleocapsid (N) in AY.104 was significantly higher compared to AY.28 (p< 0.01) and B.1.617.2 (p<0.05). Interestingly, ORF7a of AY.104 expressed significantly lower sgRNA compared to AY.28 (p < 0.0001) and B.1.617.2 (p < 0.001). (Figure 4). During the peak of the outbreak which lasted from August to September, AY.28 accounted for 128/189 (67.7%) of the infections sequenced.

**Figure 4:**
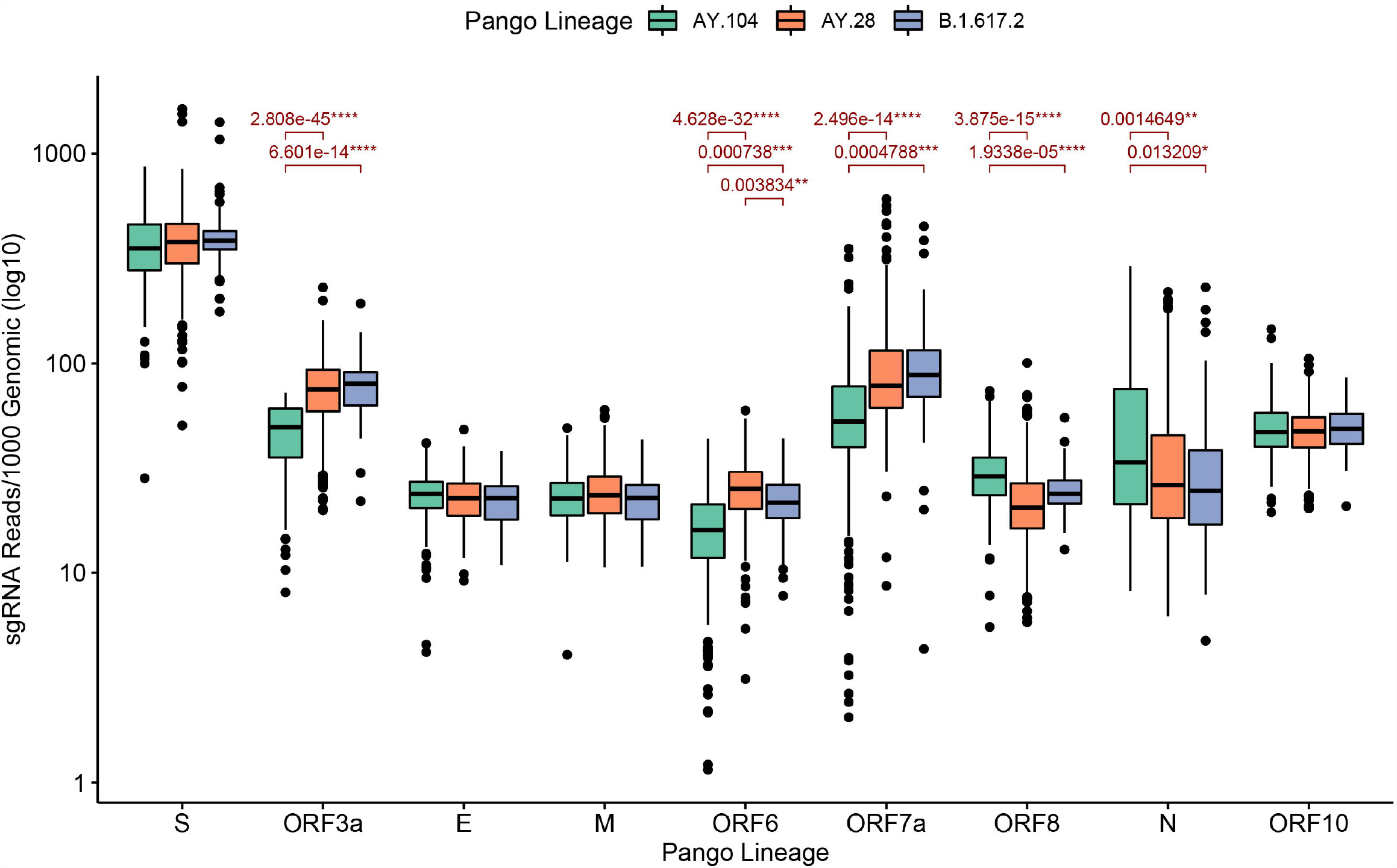
Sub-genomic RNA expression of the delta sub-lineages sequenced in Sri Lanka. Sub-genomic RNA (sgRNA) expression levels of AY.28 (n= 335), AY.104 (n= 217) and B.1.617.2 (n= 68) viruses sequenced in Sri Lanka. SgRNA reads containing the leader sequence are normalized to per 1000 genomic reads for each open reading frame (ORF) of SARS-CoV-2 using Periscope. Significant differences of mean expression levels are indicated with a p value calculated using an unpaired Wilcoxon test adjusting p-values of multiple comparisons with the Holm method (**** < 0.0001, *** < 0.001, ** < 0.01, * < 0.05).

### Clinical characteristics and vaccination status of patients infected with AY.28 and AY.104

Of the 440 individuals who were infected with the delta variant, the mean age of those infected with AY.28 was 38.39 (SD± 15.32) and AY.104 was 38.27 (SD± 17.36) and therefore was not significantly different (*p=*0.86). There was no difference in the gender of those who were infected with different delta sub-lineages (*p*= 0.39). We retrieved the clinical details of 440 patients infected with B.1.617.2 and the sub lineages from the Colombo district (Colombo Municipality Council area) and investigated if infection with different sub lineages associated with clinical disease severity. 156/160 (98%) individuals infected with AY.28 had mild infection, while 3/160 (1.88%) had moderate/severe illness and one individual succumbed to the illness. Of those who were infected with AY.104 all 63/63 (100%) patients developed mild illness. All those who were infected with B.1.617.2 18/18 (100%) also developed mild illness and there were no deaths. However, there was no significant difference between clinical disease severity in these three groups (*p=* 0.72).

Individuals were hospitalized between an average of 3 to 60 days. The mean duration of hospitalization of those infected with AY.28 was 13.91 (SD±5.88) days, for AY.104 a mean of 14.33 (SD±5.93) days and for B.1.617.2 infection a mean of 18.13 (SD±11.94) days. The duration of hospitalization was only significantly different between AY.28 and B.1.617.2 (*p=*0.0417).

In our cohort, 166/228 (72.8%) had received Sinopharm/BBIBP-CorV, while Covishield/ AZD1222 was taken by 61/228 (26.8%) individuals. Only one patient had received the Pfizer vaccine. 225/309 (72.8%) were vaccinated with at least one dose of either Sinopharm/ BBIBP-CorV, Pfizer, or the Covishield (AZD1222) vaccine while 84/309 (27.2%) were unvaccinated. Even though we observed more vaccinated people with AY.28 infection 142/309 (46%), the association was not significant (*p=*0.35). Although 178/228 (78.0%) were fully vaccinated while 50/228 (21.9%) had only one dose, we did not observe any significant association between the number of vaccine doses and infection with different sub-lineages (*p*= 0.69). Association between vaccine and sub-lineage was insignificant (*p=* 0.83).

## Discussion

In this study we have described the molecular epidemiology of the delta variant and its sub lineages during a massive outbreak in Sri Lanka, which occurred from July to November 2021. We detected the first delta variant in the community in the Colombo Municipality Council area in the third week of May 2021. This initial cluster which originated in Colombo belonged to AY.28 sub-lineage, while the first AY.104 infection was detected in the 2^nd^ week of June. By the first week of December, of the delta variants sequenced, 46.3% were of the AY.28 sub lineage, while only 33.6% were of AY.104 sub lineage. Our sequencing data from the Colombo district shows that AY.28 was the dominant variant throughout the outbreak, while AY.28 accounted for 67.7% of infections during the peak of the outbreaks (August and September). Therefore, AY.28 appears to have a fitness advantage over the parental delta variant (B.1.617.2) and AY.104, possibly due to the presence of the A222V mutation, which has been previously suggested to associate with higher transmissibility^7^. It was shown that the presence of the A222V mutation promotes an increased opening of the receptor binding domain (RBD) and slightly increases binding to ACE2 compared to the D614G SARS-CoV-2 variant^16^. However, AY.104 is also possibly more transmissible than the parental delta variant (B.1.617.2) as this only accounted for 3.39% of the sequenced variants by the first week of December.

The spike protein of AY.28 has 13 mutations (>75% prevalence) while AY.104 and the parental delta variant (B.1.617.2) have only 11 and 10 mutations respectively. Furthermore, the ORF1a of AY.28 has 7 mutations, whereas AY.104 and B.1.617.2 have 6 mutations. However, AY.28 has fewer mutations in ORF1b and nucleocapsid (N) proteins compared to the other two lineages. Unfortunately, due to the non-availability of biosafety 3 level laboratory facilities, we could not isolate these viruses and further characterize the significance of these mutations. Our analysis of clinical disease outcomes and duration of hospitalization in a sub cohort of individuals infected with AY.28, AY.104 and B.1.617.2 showed that there was no difference in the clinical disease severity or duration of hospitalization in individuals infected with these sub lineages. However, only 3 individuals in those cohort developed severe disease, while only one individual succumbed to the illness. Therefore, since only 4/440 (0.9%) individuals (all infected with AY.28) had adverse disease outcomes, the sample size is unlikely to be adequate to determine if infection with these sub lineages associate with more severe disease.

Structural proteins of coronaviruses are first transcribed into sgRNA before translation^17^. Although the presence of sgRNA per se does not indicate the presence of actively replicating virus, the relative abundance of sgRNA indicates the relative expression of different ORFs of the virus^17,18^. Many ORFs of the SARS-CoV-2 have shown to suppress interferon gene transcription, interferon production and recognition by innate immune responses, thereby inhibiting innate immune antiviral responses^19^. Although our data showed that there was no difference in the total sgRNA levels between the two sub-lineages and the parental delta, sg RNA expression was significantly higher in AY.28 for ORF3a, ORF6 and ORF7a compared to the other two lineages. All these ORFs play an important role in evading the host interferon responses by suppressing STAT1 and STAT2 phosphorylation, inhibiting STAT1 complex nuclear translocation and interacting with STING and preventing nuclear translocation of NFκβ^19-21^. Therefore, increased expression of sgRNA of ORF3a, ORF6 and ORF7a by AY.28 compared to other lineages, may associate with increased virulence due to suppression of host IFN responses. AY.104 had a higher sgRNA expression of ORF8 and the N genes compared to the other lineages. ORF8 has shown to inhibit IRF3 nuclear translocation and N protein has shown to inhibit RIG-1 signaling^19^. Therefore, both AY.28 and AY.104 had a significantly higher sgRNA expression for different ORFs compared to the parental delta variant, B.1.617.2 and thus might possibly result in more immune evasion and virulence than the parental variant. However, these sgRNA analysis data are only suggestive of such a possibility and isolation of these viruses and further studies *in vitro* and *in vivo* would be required to draw further conclusions.

The AY.28 and AY.104 delta sub lineages that originated in Sri Lanka, spread to several countries within a few weeks. AY.28 was detected in 42 countries by now, while it has predominantly been reported in USA, Japan, India and United Kingdom, reflecting the main travel destinations of Sri Lankan individuals^10^. Large divergent cluster of AY.28 seen in USA appears to be a spread of an individual case from early days of the lineage according to the most recent common ancestor (tMRCA) in late January 2021. AY.104 is currently detected in 22 countries, while again predominantly been reported in United Kingdom, India, Canada and Qatar^11^. One of the main other sub lineages reported in Sri Lanka was AY.95 (8.2%), which in thought to have originated from the Maldives^22^. Therefore, this sub lineage appears to have been introduced due to frequent travel between Sri Lanka and Maldives.

In conclusion, the massive outbreak due to the delta variant in 2021 was predominantly due to two delta sub lineages AY.28 and AY.104. These two Sri Lankan sub lineages accounted for over 80% of the sequenced delta variants in Sri Lanka from July to December, while AY.28 was the predominant sub lineage. AY.28 is possibly more transmissible than the other two lineages potentially due to the A222V mutation and significantly higher sgRNA expression was detected in ORF3a, ORF6 and ORF7a suggesting possible enhanced suppression of interferon genes compared to the parental delta variant. It would be important to further investigate the relevance of the findings by isolating these viruses, in order to understand the evolution and virulence of SARS-CoV-2 variants during this pandemic.

## Data Availability

Data is available in the manuscript and the supporting files.

## Data Availability

All data are available in the manuscript and the supporting files. The source code and data used to produce the results and analyses presented in this manuscript are available on GitHub repository https://github.com/aicbu.

## References

1 Medicine, J. H. U. a. Coronavrus Resource Centre, <https://coronavirus.jhu.edu/> (2021).

2 Butt, A. A. et al. Severity of Illness in Persons Infected With the SARS-CoV-2 Delta Variant vs Beta Variant in Qatar. JAMA Intern Med, doi:10.1001/jamainternmed.2021.7949 (2021).

3 Luo, C. H. et al. Infection with the SARS-CoV-2 Delta Variant is Associated with Higher Recovery of Infectious Virus Compared to the Alpha Variant in both Unvaccinated and Vaccinated Individuals. Clin Infect Dis, doi:10.1093/cid/ciab986 (2021).

4 Hadfield, J. et al. Nextstrain: real-time tracking of pathogen evolution. Bioinformatics 34, 4121–4123, doi:10.1093/bioinformatics/bty407 (2018).

5 Arora, P. et al. Delta variant (B.1.617.2) sublineages do not show increased neutralization resistance. Cellular & molecular immunology 18, 2557–2559, doi:10.1038/s41423-021-00772-y (2021).

6 Agency, T. U. H. S. SARS-CoV-2 variants of concern and variants under investigation in England–Technical briefing 25, 15 October 2021. (The UK Health Security Agency, 2021).

7 Hodcroft, E. B. et al. Spread of a SARS-CoV-2 variant through Europe in the summer of 2020. Nature 595, 707–712, doi:10.1038/s41586-021-03677-y (2021).

8 Lassauniere, R. et al. Neutralisation of the SARS-CoV-2 Delta variant sub-lineages AY.4.2 and B.1.617.2 with the mutation E484K by Comirnaty (BNT162b2 mRNA) vaccine-elicited sera, Denmark, 1 to 26 November 2021. Euro Surveill 26, doi:10.2807/1560-7917.ES.2021.26.49.2101059 (2021).

9 Ritchie H. O.-O. E., Beltekian D., Mathieu E., Hasell J., Macdonald B., Giattino C., Appel C., Lucas Rodés-Guirao, Rose M. (http://OurbWorldInData.org, 2021).

10 Alaa Abdel Latif, J. L. M., Manar Alkuzweny, Ginger Tsueng, Marco Cano, Emily Haag, Jerry Zhou, Mark Zeller, Emory Hufbauer, Nate Matteson, Chunlei Wu, Kristian G. Andersen, Andrew I. Su, Karthik Gangavarapu, Laura D. Hughes, and the Center for Viral Systems Biology. AY.28 Lineage Report, <https://outbreak.info/situation-reports?pango=AY.28&loc=IND&loc=GBR&loc=USA&selected> (2021).

11 Alaa Abdel Latif, J. L. M., Manar Alkuzweny, Ginger Tsueng, Marco Cano, Emily Haag, Jerry Zhou, Mark Zeller, Emory Hufbauer, Nate Matteson, Chunlei Wu, Kristian G. Andersen, Andrew I. Su, Karthik Gangavarapu, Laura D. Hughes, and the Center for Viral Systems Biology. AY.104 Lineage Report, <https://outbreak.info/situation-reports?pango=AY.104&loc=IND&loc=GBR&loc=USA&selected> (2022).

12 WHO. Clinical management of severe acute respiratory infection when novel coronavirus (2019-nCoV) infection is suspected: interim guidance. (WHO, 2020).

13 Sagulenko, P., Puller, V. & Neher, R. A. TreeTime: Maximum-likelihood phylodynamic analysis. Virus Evol 4, vex042, doi:10.1093/ve/vex042 (2018).

14 Duchene, S. et al. Temporal signal and the phylodynamic threshold of SARS-CoV-2. Virus Evol 6, veaa061, doi:10.1093/ve/veaa061 (2020).

15 Jeewandara, C. et al. Genomic and Epidemiological Analysis of SARS-CoV-2 Viruses in Sri Lanka. Frontiers in Microbiology 12, doi:10.3389/fmicb.2021.722838 (2021).

16 Ginex, T. et al. The structural role of SARS-CoV-2 genetic background in the emergence and success of spike mutations: the case of the spike A222V mutation. bioRxiv, 2021.2012.2005.471263, doi:10.1101/2021.12.05.471263 (2021).

17 Song, Z. et al. From SARS to MERS, Thrusting Coronaviruses into the Spotlight. Viruses 11, doi:10.3390/v11010059 (2019).

18 Alexandersen, S., Chamings, A. & Bhatta, T. R. SARS-CoV-2 genomic and subgenomic RNAs in diagnostic samples are not an indicator of active replication. Nat Commun 11, 6059, doi:10.1038/s41467-020-19883-7 (2020).

19 Min, Y. Q. et al. Immune evasion of SARS-CoV-2 from interferon antiviral system. Comput Struct Biotechnol J 19, 4217–4225, doi:10.1016/j.csbj.2021.07.023 (2021).

20 Xia, H. et al. Evasion of Type I Interferon by SARS-CoV-2. Cell reports 33, 108234, doi:10.1016/j.celrep.2020.108234 (2020).

21 Rui, Y. et al. Unique and complementary suppression of cGAS-STING and RNA sensing-triggered innate immune responses by SARS-CoV-2 proteins. Signal Transduct Target Ther 6, 123, doi:10.1038/s41392-021-00515-5 (2021).

22 Alaa Abdel Latif, J. L. M., Manar Alkuzweny, Ginger Tsueng, Marco Cano, Emily Haag, Jerry Zhou, Mark Zeller, Emory Hufbauer, Nate Matteson, Chunlei Wu, Kristian G. Andersen, Andrew I. Su, Karthik Gangavarapu, Laura D. Hughes, and the Center for Viral Systems Biology. AY.95 Lineage Report, <https://outbreak.info/situation-reports?pango=AY.95&loc=IND&loc=GBR&loc=USA&selected> (2022).

