## Supplementary methods for "Molecular epidemiology of AY.28 and AY.104 delta sub-lineages in Sri Lanka"

Oxford Nanopore Technology Library Preparation

The extracted RNA was quantified with NanoDrop™ 2000c Spectrophotometers (Thermo Fisher Scientific, USA). The reverse transcription was performed with LunaScript™ RT SuperMix. Then tiled PCR amplicons were generated from the SARS-CoV-2 viral cDNA with the specific primers designed by Freed et al., 2020. All the thermal cycling steps were carried out in a QuantStudio™ 5 Real-Time PCR Instrument (Applied biosystems, Singapore). The resultant 1200 bp amplicons were ligated with rapid barcodes, after ligation all the amplicons were pooled together and suspended in the equal volume of SPRI beads. The cleanup of the beads were performed with the 80% ethanol and pellet was let dried and eluted in 30 μl of elution buffer. The resultant library was quantified using the Qubit dsDNA High Sensitivity assay kit on a Qubit 4.0 instrument (Life Technologies) and 800ng of libraries in 11 μl with the addition of Rapid Adapter-F loaded in the Nanopore Mk1C.

TruSeq Stranded Total RNA Library Preparation

The starting RNA concentrations of the extracted samples were quantified using a NanoDrop™ 2000c Spectrophotometers (Thermo Fisher Scientific, USA). Ribosomal RNA was removed using biotinylated, target-specific oligos combined with Ribo-Zero rRNA removal beads. Following purification, the RNA was fragmented into small pieces using divalent cations under elevated temperature. All the thermal cycling steps were carried out in a CFX96 Deep Well, Real time System (Bio-Rad, USA). Following first and second strand cDNA synthesis 3’ ends of the blunt fragments were adenylated. Adapter ligation was carried out using IDT for Illumina- TruSeq RNA ud Indexes (Illumina, San Diego, USA). Ligated fragments were cleaned up using Agencourt AMPure XP beads (Beckman Coulter Genomics, USA) followed by DNA fragment enrichment using PCR. The libraries were quantified by using the Qubit dsDNA High Sensitivity assay kit on a Qubit 4.0 instrument (Life Technologies), and the fragment sizes (260bp) were analyzed by gel electrophoresis using a 1% gel. Each sample library was normalized to 10 nM concentration, and the normalized libraries were pooled and denatured with 0.2N NaOH. The denatured library pool was further diluted to 1.5pM and was sequenced on an Illumina NextSeq 550 platform.

Sequence Data Analysis

Samples were sequenced on Illumina Nextseq 550 platform. Resultant base calls files were demultiplexed and converted to fastq files via BaseSpace Sequence Hub (Illumina, San Diego, USA) fastq generator. Base quality of the sequenced files were checked and confirmed using fastqc[1] tool. To assess the quantity of host genomic DNA and other contaminants, Kraken 2 (version 2.0.1) Metagenomics pipeline[2] was used on the BaseSpace Sequence Hub and the abundance data were visualized using Krona[3] charts. After confirming the quantities of host DNA and viral DNA, FASTQ files were again piped through DRAGEN[4] RNA Pathogen Detection (version 3.5.14) pipeline to map reads against SARS-CoV-2 Wuhan-Hu-1 isolate (Genbank accession number: MN908947.3). The DRAGEN pipeline generated coverage plots, binary alignment files (BAM)[5] and variant call files (VCF)[6] against the accession MN908947.3. Since all four metagenomic sequences had very high depth over the SARS-CoV2 genome, consensus sequences were directly generated from the hard-filtered VCF files using bcftools (version 1.10.2)[5] consensus algorithm.

Amplicon based targeted sequencing of 150bp paired end library was performed on Illumina iSeq100 and Nextseq 550 platforms. Raw base calls of 236 samples were converted to FASTQ files using BaseSpace Sequencing Hub Fastq generator and inspected with fastqc for low quality reads. FASTQ files were aligned against SARS-CoV-2 Wuhan-Hu-1 isolate (Genbank accession number: MN908947.3) and BAM and VCF files were generated via DRAGEN RNA Pathogen Detection (version 3.5.14) pipeline. Resulting BAM files were checked using SAMTOOLS[5] (version 1.10) flagstat algorithm for alignment quality and samples with more than 86% genome coverage and moderate to low coverage depth were proceeded to obtain consensus sequences using DRAGEN pipeline. The variants were visualized and further analyzed with the VCF files alongside the BAM alignments by Interactive Genomics Viewer[7] (IGV) in order to confirm frameshift mutations.

**References**

[1] Andrews S. FastQC: A Quality Control Tool for High Throughput Sequence Data. . 2010.

[2] Wood DE, Salzberg SL. Kraken: ultrafast metagenomic sequence classification using exact alignments. Genome Biol. 2014;15:R46.

[3] Ondov BD, Bergman NH, Phillippy AM. Interactive metagenomic visualization in a Web browser. BMC Bioinformatics. 2011;12:385.

[4] Basespace.illumina.com. DRAGEN-RNA-Pathogen-Detection. Illumina; 2019.

[5] Li H, Handsaker B, Wysoker A, Fennell T, Ruan J, Homer N, et al. The Sequence Alignment/Map format and SAMtools. Bioinformatics. 2009;25:2078-9.

[6] Danecek P, Auton A, Abecasis G, Albers CA, Banks E, DePristo MA, et al. The variant call format and VCFtools. Bioinformatics. 2011;27:2156-8.

[7] Robinson JT, Thorvaldsdottir H, Winckler W, Guttman M, Lander ES, Getz G, et al. Integrative genomics viewer. Nature biotechnology. 2011;29:24-6.
